# Trends in Hepatitis C Virus Seroprevalence and Associated Risk Factors among MSM in Pakistan: Insights from a Community-Based Study

**DOI:** 10.1101/2023.09.19.23295785

**Authors:** Raza Tirmizi, Rimsha Munir, Nousheen Zaidi

**Affiliations:** Dostana Male Health Society, Pakistan; Action Research Collective, Institute of Microbiology and Molecular Genetics, University of the Punjab, Pakistan; Hormone Lab Lahore, Institute of Microbiology and Molecular Genetics, University of the Punjab, Pakistan; Cancer Biology Lab, Institute of Microbiology and Molecular Genetics, University of the Punjab, Pakistan; Cancer Research Centre, University of the Punjab, Pakistan

**Author notes:** Corresponding author: Nousheen Zaidi, University of the Punjab.

**Keywords:** HCV, Seroprevalence, MSM, HCV risk factors

## Abstract

This community-based cross-sectional study investigates the seroprevalence of the Hepatitis C Virus (HCV) and its associated demographic and behavioral risk factors among the men who have sex with men (MSM) in Pakistan. The study reveals an HCV seroprevalence of 14.86%, significantly higher than global averages for the MSM population. Notably, HCV seroprevalence was associated with age, education level, self-identified sexual orientation, and self-reported HIV status. Furthermore, the study identified several risk factors positively associated with HCV seroprevalence, including sharing personal items such as razors and toothbrushes, histories of surgery, blood transfusion, dental procedures, intravenous drug use, and therapeutic injection histories. Interestingly, a lower HCV positivity rate was observed among self-reported HIV-positive individuals, contradicting previous research. The findings underscore the need for comprehensive, targeted prevention strategies tailored to the MSM population in Pakistan. Further research is warranted to validate these findings and to understand better the complex interplay of factors contributing to HCV seroprevalence in this high-risk population.

## Introduction

Hepatitis C virus (HCV) infection, a leading cause of liver disease worldwide, manifests significant prevalence variation among different populations and locations ^1^. Multiple factors, including socioeconomic conditions ^2-4^, healthcare accessibility ^5^, and the existence of harm reduction programs ^6^, inform this epidemiological diversity. Globally, two distinct demographic groups are mainly affected by HCV infections: younger individuals aged 20-40 years and older individuals over 50 years ^7,8^.

HCV is transmitted predominantly *via* contact with contaminated blood through means such as contaminated surgical instruments, transfusion of contaminated blood or blood products, and sharing of needles or syringes ^9-11^. While the risk of transmission through sexual contact is relatively low, it is considerably higher among men who have sex with men (MSM) ^12,13^. For MSM, especially those living with the human immunodeficiency virus (HIV), unprotected sex, notably when combined with chemsex, is recognized as a potential HCV transmission route ^14,15^. Studies have consistently shown a higher prevalence of HCV infection in MSM compared to the general population ^10,16,17^.

Notably, Pakistan bears a significant burden of HCV infection, with the second-highest prevalence globally ^18^. Punjab, the most populous province in Pakistan, is known to have an exceptionally high prevalence of Hepatitis C Virus (HCV) infections. Approximately 10 million Pakistanis are estimated to be infected with HCV, with around 6.1% of these cases being active ^19,20^. Despite this high prevalence, our understanding of behavioral determinants linked to HCV infection among MSM in Pakistan, a society where marginalized groups face formidable educational and healthcare barriers, remains limited.

In the Pakistani context, it is important to note that only limited data on the seroprevalence of HCV among MSM is currently available. Furthermore, the existing data is derived from sporadic studies with small sample sizes, lacking detailed assessments of risk factors and sexual behavior patterns ^19-23^. This highlights the critical need for a comprehensive study that addresses these limitations and provides a more thorough understanding of HCV seroprevalence and its associated factors among MSM in Pakistan.

By conducting a robust investigation encompassing a larger sample size and a detailed assessment of risk factors and sexual behaviors, our study aims to fill this knowledge gap and contribute to evidence-based interventions and strategies for HCV prevention and control among MSM in Pakistan. Our study overcame the challenges of researching disease prevalence in marginalized communities, such as Pakistan’s MSM population, by collaborating with an organization *Dostana Male Health Society*. Established in 2012 under the Societies Registration Act 1860, *Dostana* specializes in HIV prevention and community empowerment for at-risk and marginalized populations. Through our partnership with *Dostana*, we successfully engaged with the MSM community, addressing their unique barriers and conducting the study effectively. This collaborative approach aims to enhance our understanding of HCV infection dynamics among MSM in Pakistan, thus informing strategies to combat the HCV infection epidemic in this uniquely challenging social and healthcare setting.

## Methodology

### Research Design and Participant Selection

This cross-sectional study determines the seroprevalence of the Hepatitis C Virus (HCV) among MSM in Pakistan. A total of 501 participants were included in this study. The inclusion criteria stipulated that participants must be MSM aged 15 years or older and possess proficiency in Punjabi or Urdu. Participants were recruited through the *Dostana Male Health Society*, a recognized society that addresses key populations’ healthcare concerns. Due to Pakistan’s societal and cultural barriers, engaging with individuals in these marginalized communities is challenging. As a solution, community members were educated and guided to gather data from the MSM population enlisted through *Dostana*.

The data collection process involved structured interviews. After the necessary training, data enumerators administered the structured questionnaires through one-on-one interviews. This method ensured that the process was consistent, comprehensive, and aimed at minimizing potential bias or misinterpretation of the questions. The data were collected on demographic characteristics, common risk factors, and history of HIV.

### Ethical Considerations

The study protocol is reviewed and approved by the Bioethics Committee of the Cancer Research Centre, University of the Punjab. All participants provided informed written consent before participating in the study. The study is conducted following the WMA Declaration of Helsinki-Ethics principles for research involving human subjects.

### Serological Testing

Blood samples are collected from participants for serological testing. The presence of HCV antibodies is determined using the SD BIOLINE HCV Hepatitis C Virus Antibody Test. All tests are conducted in a certified laboratory following standard procedures.

### Statistical Analysis

Descriptive statistics are used to summarize the demographic characteristics of the participants and the Prevalence of HCV. All analyses are performed using MS Excel.

## Results

### Demographic Characteristics and HCV Seroprevalence

For the presented work, 501 MSM participants were recruited to assess the seroprevalence of HCV. The data on the demographic characteristics were recorded through a structured interview by our survey enumerators. An analysis of the demographic characteristics within the study population revealed several striking patterns. Firstly, the age distribution highlighted a substantial representation of younger individuals, with participants aged 18-35 comprising the majority (∼75%) of the total sample (**Table 1**). Additionally, an intriguing observation emerged regarding educational attainment, as a significant proportion of participants reported no formal education (∼20%) or had completed only five years (∼28%) or less(**Table 1**). Our results indicate that HCV was positive in 14.86% of the study population (**Figure 1a**)

**Table 1:** Demographics of the study-population.

| **Table 1: Demographics of the study-population** |  |
| --- | --- |
| **Demographic Criteria** | **Number of Participants** |
| **Total Population (n)** | 501 |
| **Age Groups (n)** |  |
| <18 | 7 |
| 18-25 | 148 |
| 26-35 | 230 |
| 36-45 | 84 |
| >45 | 32 |
| **Years of Formal Education** |  |
| 0 | 103 |
| ≤5 | 139 |
| 6-10 | 152 |
| >10 | 107 |
| **Sexual Orientation** |  |
| Hetero | 30 |
| Homo | 269 |
| Bi | 191 |
| ND | 11 |
| **HIV Status** |  |
| HIV +ve | 212 |
| HIV -ve | 289 |

**Figure 1:**
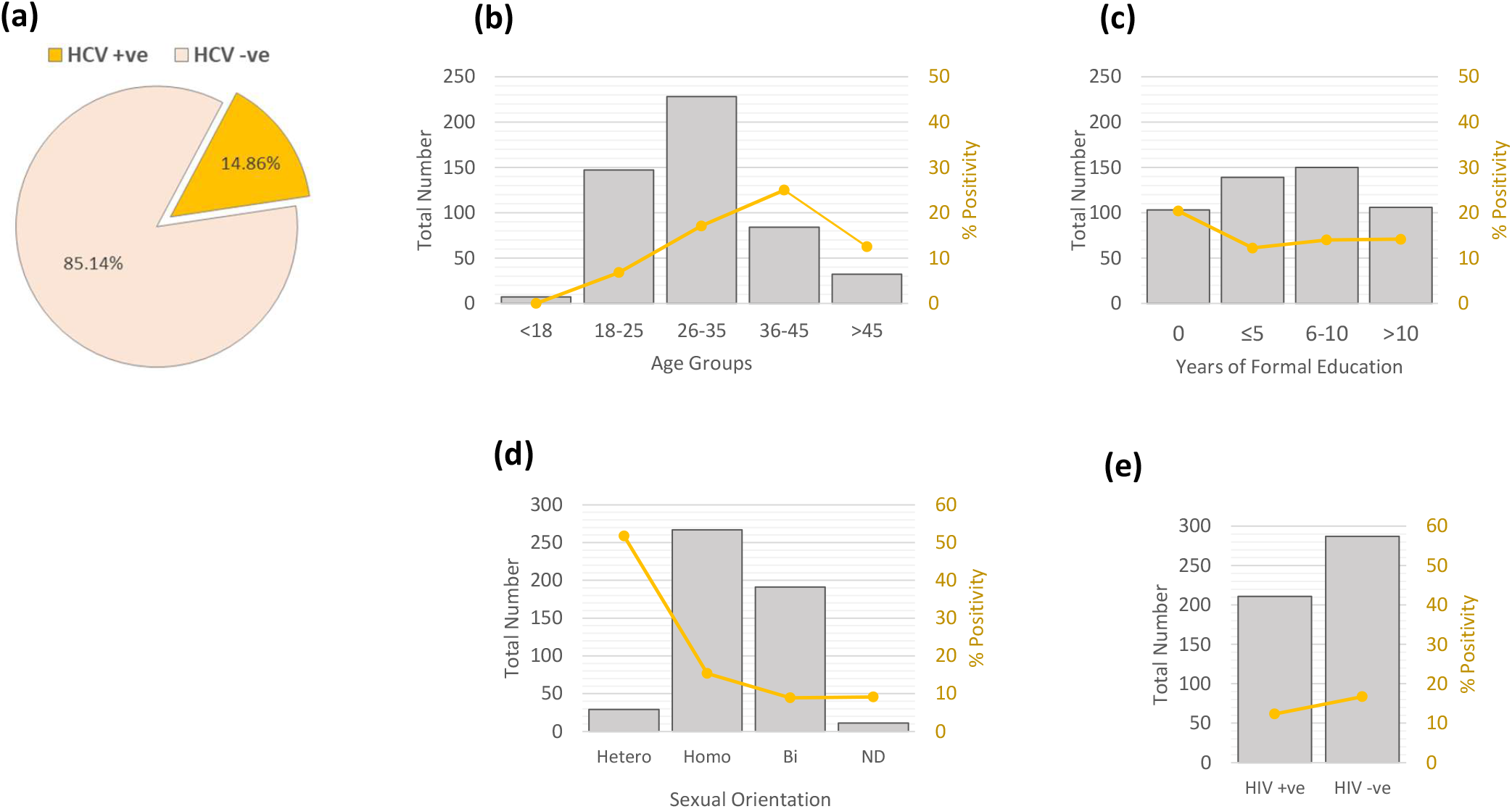
Prevalence of HCV seropositivity and associated demographic risk factors in Pakistani MSM. **(a)** A pie-chart displaying seroprevalence of HCV in our MSM population (n=501). The total number of MSM participants in each **(b)** Age **(c)** Education **(d)** Sexual orientation and **(e)** Self-reported HIV status groups are plotted on *left y-axis* and represented as vertical *grey bars* and HCV seropositivity percentage is plotted on *right y-axis* and represented as a *yellow stacked line*.

### Association of HCV seroprevalence with selected demographic characteristics

We explored the relationships between demographic characteristics and HCV seroprevalence in our MSM population. We observed a noticeable trend of HCV positivity rates with varying age groups (**Figure 1b**). From age 18, the HCV positivity increased steadily up to the age group of 36-45 years, which reported the highest HCV prevalence at 25%. Surprisingly, the rate decreased to 12.5% in respondents over 45.

We found an inverse relationship between years of formal education and the HCV positivity rate. The group with no formal education had the highest HCV positivity at 20.39%. However, HCV positivity remained relatively consistent as education levels increased (**Figure 1c**).

Our data demonstrated variability in HCV positivity rates based on self-reported sexual orientation. Heterosexual respondents had a markedly high HCV positivity rate of 51.7%, significantly higher than in homo-, bi-, or undisclosed sexual orientation respondents, who reported HCV positivity rates of 15.3%, 8.9%, and 9.0%, respectively (**Figure 1d**).

Further, we examined the interplay between HIV status and HCV positivity. Interestingly, a lower HCV positivity rate (12.3%) was observed among HIV-positive individuals compared to their HIV-negative counterparts (16.7%) (**Figure 1e**).

### Association of HCV seroprevalence with common risk factors

Next, we analyzed the relationship between HCV seropositivity and various commonly identified risk factors within the MSM population. The risk factors assessed were: utilization of barber services, sharing of razors or toothbrushes, surgical history, history of blood transfusion, dental procedures, intravenous and oral drug abuse, syringe reuse, and therapeutic injection history (**Figure 2**).

**Figure 2:**
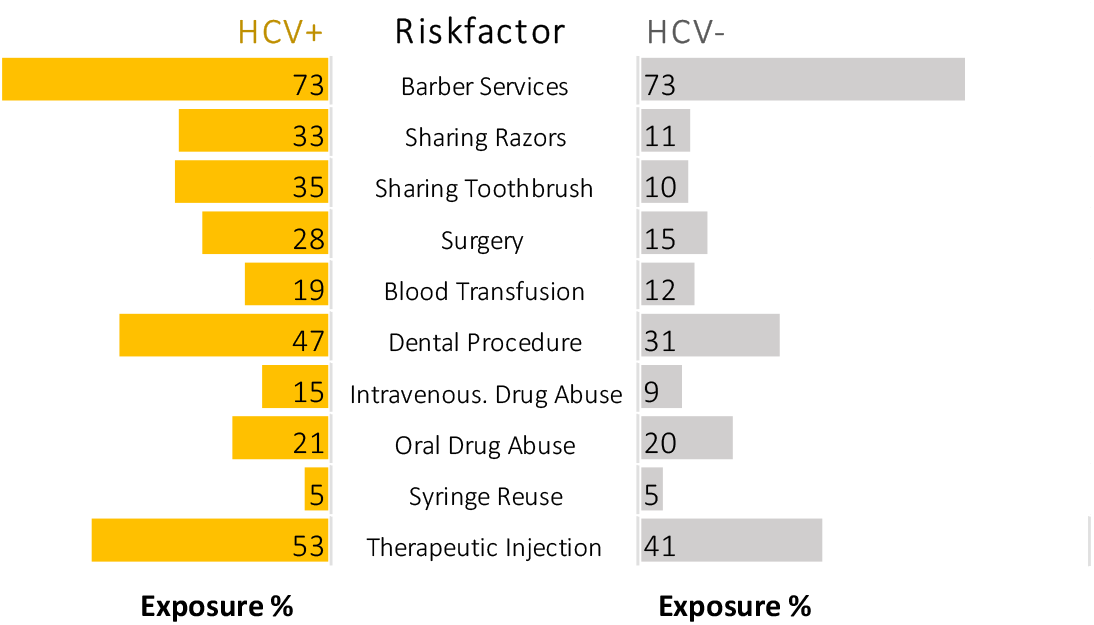
Exposure to selected behavioral risk factors in seropositive and seronegative MSM population. A *butterfly graph* displaying exposure percentage of different behavioral risk factors in seropositive and seronegative MSM population.

The prevalence of each risk factor was compared between the HCV-positive and HCV-negative groups. Our data indicated a high percentage of individuals that use barber services in both the HCV-positive (73%) and HCV-negative groups (73%). Similarly, we observed comparable proportions of syringe reuse and oral drug abuse between the HCV-positive and HCV-negative groups.

However, several risk factors were markedly more prevalent in the HCV-positive group compared to the HCV-negative group. The sharing of razors was reported by 33% of HCV-positive individuals compared to 11% in the HCV-negative group. Similarly, sharing toothbrushes was significantly more prevalent in the HCV-positive group (35%) than in the HCV-negative group (10%).

Additionally, histories of surgery and blood transfusion were more common in the HCV-positive group (28% and 19%, respectively) compared to the HCV-negative group (15% and 12%, respectively). Dental procedure histories, intravenous drug abuse, and therapeutic injection histories were also higher in the HCV-positive group.

### Association of HCV seroprevalence with sexual risk behaviors

Next, we investigated the association between sexual risk behaviors and HCV seroprevalence. The dataset included the percentage of HCV seropositivity for different sexual risk behaviors. Regarding the number of sex partners, individuals with less than 5 partners had an HCV positivity rate of 11%, while those with 5-20 partners exhibited a higher rate of 18%. Participants with more than 20 partners had a lower HCV positivity rate of 9% (**Figure 3**).

**Figure 3:**
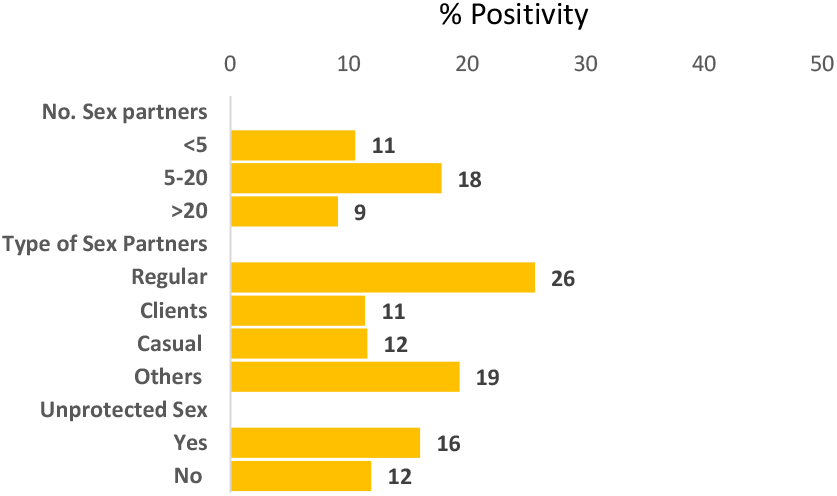
Association between HCV seropositivity and sexual risk behaviors in MSM. A *bar graph* displaying seropositivity percentage in MSM population engaging in different sexual risk behaviors.

Examining the type of sex partners, individuals involved with regular partners showed the highest HCV positivity rate at 26%. Those engaged with clients and casual partners had rates of 11% and 12%, respectively. Individuals involved with other types of partners had an HCV positivity rate of 19%.

Evaluating unprotected sex, participants who reported engaging in unprotected sexual activities had an HCV positivity rate of 16%, whereas those who practiced protected sex had a rate of 12%. These findings highlight the varying degrees of HCV positivity among individuals based on their sexual risk behaviors, emphasizing the importance of promoting safe sexual practices and tailored interventions to reduce HCV transmission.

## Discussion

In this study, we sought to explore the associations between Hepatitis C Virus (HCV) seroprevalence and selected demographic characteristics, risk factors, and sexual behaviors among the men who have sex with men (MSM) population in Pakistan. Our findings revealed an HCV seroprevalence of 14.86%, significantly exceeding the global pooled seroprevalence for HCV infection in MSM, which stands at 3.4% (95% CI 2.8– 4.0) ^24^. This discrepancy underscores the unique epidemiological landscape of HCV infection in Pakistan’s MSM population.

We found that HCV positivity increased steadily for our population from age 18 to age 36-45 years, which reported the highest HCV prevalence. However, the rate decreased in respondents over the age of 45. This pattern could be attributed to age-related risk-taking behavior or a generational difference in awareness about HCV ^25,26^, which warrants further investigation. Moreover, the increase in HCV positivity up to 36-45 years could be due to cumulative exposure to risk factors over time ^27^. The decrease in positivity rates in respondents over 45 might be due to survivor bias, where those who are infected and do not receive treatment may not survive to older ages.

Our analysis revealed a notably high HCV positivity rate among self-identified heterosexual participants within the MSM group. This observation, while compelling, should be interpreted with caution due to the small sample size of this subgroup (n=30), which limits the generalizability of this finding. With smaller sample sizes, a few cases of HCV positivity can significantly affect the overall percentage, leading to potentially inflated estimates of HCV prevalence in that group. Moreover, smaller sample sizes are generally associated with greater uncertainty and less statistical power, making it difficult to draw definitive conclusions. This subgroup’s high HCV positivity rate could be attributed to undisclosed or stigmatized same-sex behaviors, but further research is necessary to substantiate this hypothesis. Future studies with larger sample sizes are warranted to validate this finding and to delve deeper into the potential underlying factors contributing to this elevated HCV positivity rate among self-identified heterosexual MSM.

Our study revealed a lower HCV positivity rate among HIV-positive individuals than their HIV-negative counterparts. This finding contrasts previous research, which typically reports increased rates of sexually transmitted HCV infection among the HIV-positive MSM population ^28,29^. This discrepancy may be attributed to several factors. One possible explanation is that individuals living with HIV, due to their regular engagement with healthcare services, may have more frequent screening for HCV, better adherence to treatment, and more robust implementation of prevention strategies, which could contribute to a lower HCV prevalence in this group. However, it’s also crucial to note that HIV seroprevalence was not directly tested in this study. Instead, we relied on self-reported HIV status, which could introduce bias and skew the results. Individuals may underreport their HIV status due to stigma or lack of awareness, leading to an underestimation of HCV positivity among the HIV-positive population. While these explanations are plausible, they remain speculative, and further research is needed to understand this counterintuitive finding fully.

Our study identified several risk factors that were positively associated with HCV seroprevalence among the MSM population in Pakistan. Sharing personal items like razors and toothbrushes was significantly more prevalent among HCV-positive individuals. Histories of surgery, blood transfusion, and dental procedures were more common among HCV-positive individuals. Intravenous drug use and therapeutic injection histories were also higher among HCV-positive individuals. These findings suggest that sharing personal hygiene items, histories of medical procedures, and practices associated with drug use may contribute to the higher prevalence of HCV infection among MSM individuals.

This study also examined the relationship between sexual risk behaviors and HCV seroprevalence. Our analysis involved assessing the percentage of HCV positivity for different sexual risk behaviors. Among individuals with varying numbers of sex partners, those with less than 5 partners had an HCV positivity rate of 11%. In contrast, individuals with 5-20 partners exhibited a higher HCV positivity rate of 18%, indicating an increased risk of HCV transmission with a higher number of partners. Interestingly, individuals with more than 20 partners had a lower HCV positivity rate of 9%, suggesting a potential decrease in risk beyond a certain threshold of sexual partners.

When considering the type of sex partners, individuals involved with regular partners had the highest HCV positivity rate at 26%, indicating a heightened risk of HCV transmission within stable sexual relationships. The higher HCV positivity rate among individuals involved with regular sex partners could be attributed to factors such as longer-term sexual relationships, increased likelihood of sharing needles or drug-related behaviors, or a higher prevalence of HCV infection within certain social networks.

Those engaged with clients and casual partners had comparatively lower seropositivity rates of 11% and 11%, respectively. This lower seroprevalence be influenced by factors such as commercial sex work practices, the use of protective measures during sexual encounters, or lower prevalence of HCV infection within these specific population.

In conclusion, our study sheds light on the diverse factors associated with HCV prevalence among MSM and underlines the need for comprehensive, targeted prevention strategies. More research is necessary to verify our findings, particularly those related to age, education level, and sexual behavior. Our study contributes valuable insights that can guide the design of intervention programs, emphasizing education, safe practices, and increased healthcare engagement for this high-risk population.

## Data availability

The datasets supporting the conclusions of this article are included within the article.

## Consent for Publication

Not applicable

## References

1 Stanaway, J. D. et al. The global burden of viral hepatitis from 1990 to 2013: findings from the Global Burden of Disease Study 2013. Lancet 388, 1081–1088, doi:10.1016/s0140-6736(16)30579-7 (2016).

2 Pérez, C. M., Suárez, E., Torres, E. A., Román, K. & Colón, V. Seroprevalence of hepatitis C virus and associated risk behaviours: a population-based study in San Juan, Puerto Rico. International journal of epidemiology 34, 593–599 (2005).

3 Dalgard, O., Jeansson, S., Skaug, K., Raknerud, N. & Bell, H. Hepatitis C in the general adult population of Oslo: prevalence and clinical spectrum. Scandinavian journal of gastroenterology 38, 864–870 (2003).

4 Armstrong, G. L. et al. The prevalence of hepatitis C virus infection in the United States, 1999 through 2002. Annals of internal medicine 144, 705–714 (2006).

5 Astell-Burt, T., Flowerdew, R., Boyle, P. J. & Dillon, J. F. Does geographic access to primary healthcare influence the detection of hepatitis C? Social Science & Medicine 72, 1472–1481 (2011).

6 Papatheodoridis, G. & Hatzakis, A. Public health issues of hepatitis C virus infection. Best practice & research Clinical gastroenterology 26, 371–380 (2012).

7 Manns, M. P. et al. Hepatitis C virus infection. Nature Reviews Disease Primers 3, 17006, doi:10.1038/nrdp.2017.6 (2017).

8 Vanhommerig, J. W. et al. Risk Factors for Sexual Transmission of Hepatitis C Virus Among Human Immunodeficiency Virus-Infected Men Who Have Sex With Men: A Case-Control Study. Open Forum Infect Dis 2, ofv115, doi:10.1093/ofid/ofv115 (2015).

9 Alaei, K., Sarwar, M. & Alaei, A. The urgency to mitigate the spread of hepatitis C in Pakistan through blood transfusion reform. International Journal of Health Policy and Management 7, 207 (2018).

10 Midgard, H. et al. HCV epidemiology in high-risk groups and the risk of reinfection. Journal of hepatology 65, S33–S45 (2016).

11 Ramachandran, S. et al. A large HCV transmission network enabled a fast-growing HIV outbreak in rural Indiana, 2015. EBioMedicine 37, 374–381 (2018).

12 Chan, D. P., Sun, H.-Y., Wong, H. T., Lee, S.-S. & Hung, C.-C. Sexually acquired hepatitis C virus infection: a review. International Journal of Infectious Diseases 49, 47–58 (2016).

13 Jin, F., Matthews, G. V. & Grulich, A. E. Sexual transmission of hepatitis C virus among gay and bisexual men: a systematic review. Sexual health 14, 28–41 (2016).

14 Hoornenborg, E. et al. High incidence of HCV in HIV-negative men who have sex with men using pre-exposure prophylaxis. J Hepatol 72, 855–864, doi:10.1016/j.jhep.2019.11.022 (2020).

15 Platt, L. et al. Prevalence and burden of HCV co-infection in people living with HIV: a global systematic review and meta-analysis. Lancet Infect Dis 16, 797–808, doi:10.1016/s1473-3099(15)00485-5 (2016).

16 Jachs, M. et al. Outcomes of an HCV elimination program targeting the Viennese MSM population. Wiener klinische Wochenschrift 133, 635–640 (2021).

17 Han, R., Zhou, J., François, C. & Toumi, M. Prevalence of hepatitis C infection among the general population and high-risk groups in the EU/EEA: a systematic review update. BMC infectious diseases 19, 1–14 (2019).

18 Khan, A. et al. Molecular epidemiology and genotype distribution of hepatitis C in Pakistan; a multicenter cross-sectional study. Infection, Genetics and Evolution 84, 104372 (2020).

19 Idrees, M., Lal, A., Naseem, M. & Khalid, M. High prevalence of hepatitis C virus infection in the largest province of Pakistan. Journal of digestive diseases 9, 95–103 (2008).

20 Mahmud, S., Al Kanaani, Z. & Abu-Raddad, L. J. Characterization of the hepatitis C virus epidemic in Pakistan. BMC infectious diseases 19, 1–11 (2019).

21 Akhtar, H. et al. Prevalence of hepatitis B and hepatitis C Virus infections among male to female (MFT) transgenders in Rawalpindi (Pakistan). Advancements in Life Sciences 5, 46–55 (2018).

22 Zahra, A., Saleem, M. A., Javed, H. & Khan, M. A. U. Prevalence of HCV-HIV Co-Infection with Intravenous Drug Users in Central Punjab, Pakistan. Pak. J. Zool 54, 2003–2500 (2021).

23 Khanani, M. R., Somani, M., Khan, S., Naseeb, S. & Ali, S. H. Prevalence of single, double, and triple infections of HIV, HCV and HBV among the MSM community in Pakistan. Journal of Infection 61, 507–509 (2010).

24 Zheng, Y. et al. Global burden and changing trend of hepatitis C virus infection in HIV-Positive and HIV-Negative MSM: a systematic review and meta-analysis. Frontiers in Medicine 8, 774793 (2021).

25 Clipman, S. J. et al. Prevalence and phylogenetic characterization of hepatitis C virus among Indian men who have sex with men: Limited evidence for sexual transmission. The Journal of infectious diseases 221, 1875–1883 (2020).

26 Jin, F. et al. Prevalence and incidence of hepatitis C virus infection in men who have sex with men: a systematic review and meta-analysis. The Lancet Gastroenterology & Hepatology 6, 39–56 (2021).

27 Tseng, Y.-T. et al. Seroprevalence of hepatitis virus infection in men who have sex with men aged 18–40 years in Taiwan. Journal of the Formosan Medical Association 111, 431–438 (2012).

28 Hagan, H., Jordan, A. E., Neurer, J. & Cleland, C. M. Incidence of sexually transmitted hepatitis C virus infection in HIV-positive men who have sex with men. Aids 29, 2335–2345, doi:10.1097/qad.0000000000000834 (2015).

29 van Santen, D. K. et al. Lack of decline in hepatitis C virus incidence among HIV-positive men who have sex with men during 1990-2014. J Hepatol 67, 255–262, doi:10.1016/j.jhep.2017.03.038 (2017).

